# Accounting for external factors and early intervention adoption in the design and analysis of stepped-wedge designs: Application to a proposed study design to reduce opioid-related mortality

**DOI:** 10.1101/2020.07.26.20162297

**Authors:** Lior Rennert, Moonseong Heo, Alain H. Litwin, Victor De Gruttola

## Abstract

**Background:** Stepped-wedge designs (SWDs) are currently being used to investigate interventions to reduce opioid overdose deaths in communities located in several states. However, these interventions are competing with external factors such as newly initiated public policies limiting opioid prescriptions, media awareness campaigns, and social distancing orders due to the COVID-19 pandemic. Furthermore, control communities may prematurely adopt components of the proposed intervention as they become widely available. These types of events induce confounding of the intervention effect by time. Such confounding is a well-known limitation of SWDs; a common approach to adjusting for it makes use of a mixed effects modeling framework that includes both fixed and random effects for time. However, these models have several shortcomings when multiple confounding factors are present.

**Methods:** We discuss the limitations of existing methods based on mixed effects models in the context of proposed SWDs to investigate interventions intended to reduce mortality associated with the opioid epidemic, and propose solutions to accommodate deviations from assumptions that underlie these models. We conduct an extensive simulation study of anticipated data from SWD trials targeting the current opioid epidemic in order to examine the performance of these models under different sources of confounding. We specifically examine the impact of factors external to the study and premature adoption of intervention components.

**Results:** When only external factors are present, our simulation studies show that commonly used mixed effects models can result in unbiased estimates of the intervention effect, but have inflated Type 1 error and result in under coverage of confidence intervals. These models are severely biased when confounding factors differentially impact intervention and control clusters; premature adoption of intervention components is an example of this scenario. In these scenarios, models that incorporate fixed intervention-by-time interaction terms and an unstructured covariance for the intervention-by-cluster-by-time random effects result in unbiased estimates of the intervention effect, reach nominal confidence interval coverage, and preserve Type 1 error, but may reduce power.

**Conclusions:** The incorporation of fixed and random time effects in mixed effects models require certain assumptions about the impact of confounding by time in SWD. Violations of these assumptions can result in severe bias of the intervention effect estimate, under coverage of confidence intervals, and inflated Type 1 error. Since model choice has considerable impact on study power as well as validity of results, careful consideration needs to be given to choosing an appropriate model that takes into account potential confounding factors.

## 1. Introduction

Stepped wedge designs (SWD) are a uni-directional crossover design in which clusters switch from the control to the intervention condition at varying time points. The first phase is usually a baseline period in which no clusters receive intervention. During the second phase, clusters are randomly assigned to intervention at pre-selected time points until all clusters receive the intervention. The third phase corresponds to the follow-up period in which all clusters receive the intervention. An example of a SWD is provided in Figure 1 and discussed in Section 1.1.

**Figure 1.**
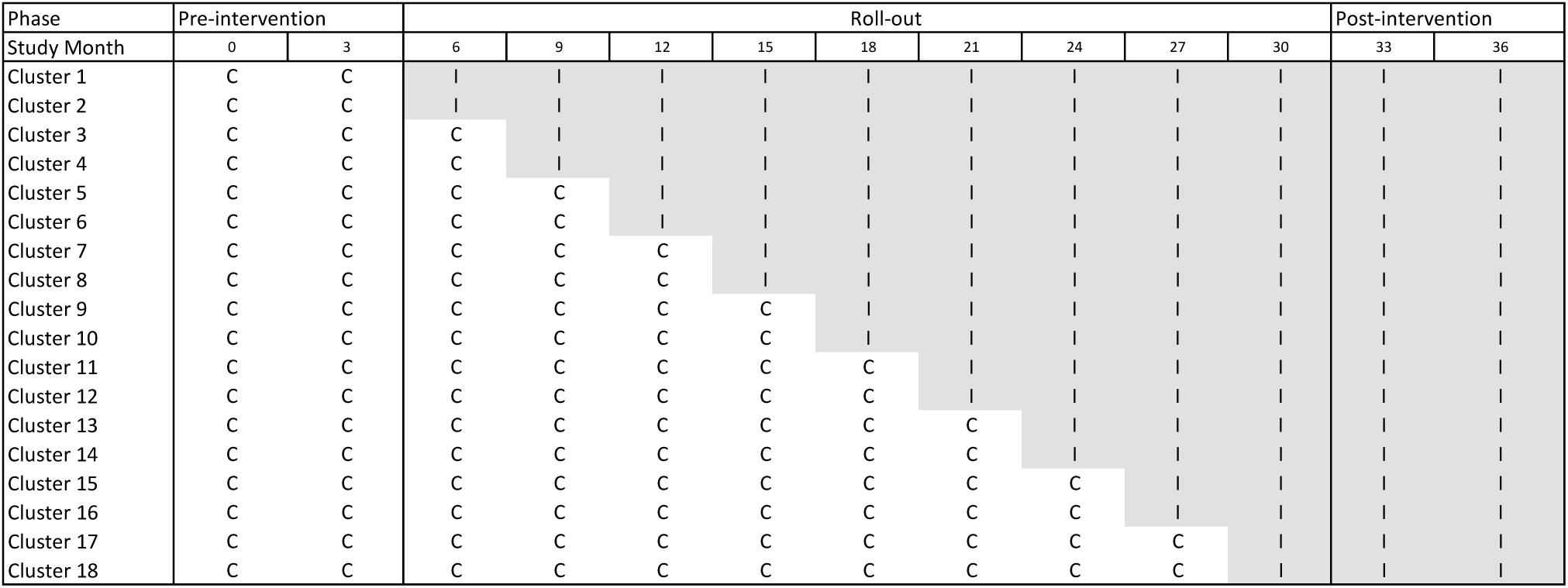
Proposed SWD for 18 South Carolina communities. ‘C’ indicates cluster receives control and ‘I’ indicates cluster receives intervention. All clusters are in the control condition during the pre-intervention phase (months 0 through 6). During the roll-out phase (months 6 through 33), two clusters crossover to the intervention condition at the beginning of each time period. In the follow-up phase (months 33-39), all clusters receive the intervention.

SWDs can be useful in public health settings where rolling out the intervention to all clusters at once is infeasible; they also ensure that all clusters in the study eventually receive the intervention.^1, 2^ The SWD is particularly suitable for implementing and evaluating complex health interventions.^3–5^ The current COVID-19 pandemic and opioid epidemic provide settings in which such designs may prove useful; effective combinations of interventions are needed as soon as possible, but rolling them out to each community in a short time period may not be feasible. In this paper, we will focus on recently proposed SWDs to combat the opioid epidemic. In 2019, the National Institute on Drug Abuse (NIDA) awarded Kentucky and Ohio roughly $100 million each to implement an integrated set of interventions in high-risk communities using a SWD, with the objective of reducing opioid overdose deaths by 40% over a three-year period.^6–9^

Like most clinical trial designs, SWDs may be impacted by external factors that influence the primary outcomes. The term “rising tide” has been used to describe the situation when there is a drift towards improvement in the outcomes due to factors external to the study.^10^ A rising tide may be seen in the current opioid epidemic, where the severity of this crisis has led to new public policies, media awareness campaigns, and external interventions that will improve outcomes in concurrence with any proposed interventions. For example, public policies were implemented to limit opioid prescriptions in both Kentucky and Ohio in the summer of 2017.^11^ In addition, the Center for Disease Control and Prevention launched intensive media awareness campaigns on the dangers of opioids in these states in September of 2017.^12^ In the current COVID-19 pandemic, interventions rolled out at the community level will compete with social distancing measures, novel treatments, and other interventions aimed at improving patient care. Such external factors may confound the intervention effect estimate in SWDs. This occurs when (i) factors external to the study (e.g., new public health policy) influence the primary outcomes over time, and (ii) the proportion of communities exposed to the intervention also increases with calendar time. This situation has been referred to as confounding by calendar time.^2, 13^

In certain scenarios, confounding factors may differentially impact intervention and control communities. For example, while all communities may be exposed to mandated state-wide policies and media awareness campaigns, control communities may choose to prematurely adopt components of the intervention prior to the scheduled roll-out time, should they become available. The latter scenario may arise from external forces, such as implementation of intervention components in control communities by policymakers, or easy access to intervention components by participants in control communities. Therefore, premature adoption of intervention components, which is a version of cluster treatment non-compliance, can also be viewed as a special case of confounding by time which impacts control communities only.

Failure to account for confounding by time in these settings can result in severely biased intervention effect estimates,^2, 3, 13–19^ and may lead to both Type 1 and Type 2 errors. Hussey and Hughes suggested the use of mixed effects models for analyzing data from SWD.^16^ To account for potential confounding by time, they recommend the incorporation of fixed time effects. However, models that incorporate secular trends common to all clusters may not be appropriate in this setting, because only some of the communities may be exposed to the external factors. Furthermore, the impact of these external factors on the outcome will likely differ in each community.

For these reasons, Girling and Hemming, Hooper et al., and Hemming et al. have suggested incorporating random cluster-by-time effects into the mixed effects models.^13, 17, 20^ This class of models captures confounding by time through these random effects. This is especially useful when the timing and level of exposure to confounding factors are unknown. However, these models implicitly assume that the random effect variance is the same for all clusters across all time points, and that the random time effects are independent within each cluster. Kazca et al. propose models with more general within-cluster correlation structures.^21^ These models are more applicable to the scenarios discussed here, since outcomes within a community are more likely to be similar for time periods before or after exposure to confounding factors. Furthermore, since exposure to confounding factors may increase with time, variation in the outcome may change with time as well.

The models described above assume that the random effects are identically distributed across clusters. This assumption may not appropriate in settings where the impact of confounding factors systematically differs between intervention and control communities. Kazca et al showed that the random effect covariance structure can have an important impact on the sample size and power,^21^ and that misspecification of this covariance structure can lead to biased estimates of the intervention effect.^22^

In this paper, we discuss confounding in the context of a proposed SWD to reduce opioid-related mortality in South Carolina, and show how trial conclusions can differ based on the choice of the fixed time effects and random effect covariance structure in mixed effects models. Our objectives are to (1) consider the implications of assumptions regarding random effects in mixed models in the setting of confounding by time– with a focus on our motivating example; (2) extend these models to accommodate different sources of confounding by time; (3) conduct an extensive simulation study to examine the performance of mixed effects models under different model assumptions regarding fixed and random time effects. Performance is assessed based on the bias, confidence interval coverage, power, and Type 1 error of the intervention effect estimate. An important component of our simulation study is the generation of latent confounding factors from different distributions than that assumed by the model, i.e. normal distribution for random effects. This allows for investigation of the robustness of our methods to violations of model assumptions.

The outline of this paper is as follows. Section 1.1 introduces the motivating example. In Section 2, we examine existing mixed effects models for SWD, discuss their limitations, and introduce alternative models that improve robustness to confounding by time. Section 3 conducts an extensive simulation study to examine the adequacy of these mixed-effects models under different scenarios of confounding by time. Discussion, extensions, and concluding remarks are provided in Section 4.

### 1.1. Motivating example: HEALing Communities Study

The HEALing Communities Study: Developing and Testing an Integrated Approach to Address the Opioid Crisis^7^ is used as an example to illustrate confounding by time in SWD. The purpose of this initiative was to develop and integrate a set of evidence based interventions using cluster randomized trials to reduce opioid overdose fatalities by 40% over a 3-year period in states nationwide. In response to this research funding announcement, our research team proposed a SWD to implement a comprehensive external facilitation intervention in 18 of South Carolina’s counties that were hit hardest by the opioid crisis.

The proposed SWD design is displayed in Figure 1. Each cluster consists of all individuals in a given county. Each time interval corresponds to a 3-month study period. The pre-intervention phase consists of two time periods between (study) month 0 and month 6, in which all clusters are in the control condition. The roll-out phase consists of nine time periods between months 6 through 33, where two clusters are randomly assigned to receive the intervention at the beginning of each time period. The post-intervention phase, in which all clusters receive the intervention, occurs between months 33 through 39. Data pertaining to the communities considered in our proposal are provided in Supplementary Table 1. The outcome, recorded at the end of each time period, consists of the total number of opioid overdose deaths in each cluster during the 3-month time interval.

The proposed external facilitation intervention included the following components: 1) Integration of screening, intervention, and referral to treatment within health care settings; 2) implementation of programs and providers prescribing medication-assisted treatment and linkage to such treatment for people with opioid use disorder (OUD); 3) implementation of evidence-based school and community-based OUD prevention programs; and 4) increasing availability and use of naloxone by first responders and the community. Given the amount of effort and resources currently directed to the fight against the opioid epidemic, there is potential for other events to affect the outcome. For example, in 2018 Executive Order No. 2017-43 was passed in South Carolina; this order set a 5-day limit for certain opioid prescriptions.^23^ Also in 2018, the South Carolina Division of Alcohol and Other Drug Abuse Services rolled out a media campaign to raise community awareness of opioid addiction.^24^ This campaign included digital, social, and traditional media tactics, and was intended to cover all counties in South Carolina. These external factors are part of a rising tide of interventions and are expected to reduce the opioid-related death count over time. Failing to account for this rising tide in an analysis will cause an upward bias in the estimation of the intervention effect. Similarly, an influx of synthetic opioids into the population will likely be associated with an increase in the death rate over time. A failure to account for this will cause underestimation of the intervention effect and may lead to Type II error. Another possibility is that control communities prematurely adopt components of the intervention before the scheduled roll-out time. For example, the opioid overdose reversal medication naloxone may become widely available in control communities prior to the scheduled roll-out time. Therefore premature adoption of a successful intervention will improve the outcome in control communities and attenuate the estimate of the intervention effect if unaccounted for.

## 2. Methods

We first introduce some notation. We denote *Y*_*ij*_ as the summary measure of the outcome for cluster *i* during time period *j, i* = 1, …, *N* and *j* = 1, …, *n*, where *N* denotes the number of clusters and *n* denotes the number of time periods in the study, which are assumed to be common to all clusters. In the motivating example, *Y*_*ij*_ is an aggregate count of opioid deaths in county *i* between months *j* and *j* + 1. Using summary measures for the outcome in each cluster has implications when estimating the intervention effect under certain distributional assumptions for the outcome. We discuss this further in Section 2.4.

We assume that the expected outcomes, denoted by *µ*_*ij*_ = *E*[*Y*_*ij*_], come from a generalized linear mixed effects model (GLMM) with link function *g*. In the motivating example we consider *µ*_*ij*_ = *E*[*Y*_*ij*_*/O*_*i*_], where *Y*_*ij*_ assumes a Poisson distribution (*g* = log link), and *O*_*i*_ is an offset for the population size of cluster *i*. We also assume that all clusters receive the full intervention effect immediately after the scheduled implementation, i.e., that the intervention effect does not change with time. We denote the intervention effect by *θ*, and set the corresponding design matrix *X*_*ij*_ = 1 if clusters *I* receives intervention at time *j*, and 0 otherwise.

### 2.1. Basic model

To adjust for confounding by calendar time, Hussey and Hughes recommended incorporation of a time effect in the GLMM:^16^

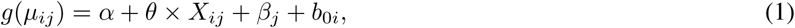

where *α* is the intercept, *θ* is the intervention effect, *β*_*j*_ is the discrete time effect, and 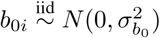 for *i* = 1, …, *N* are the random intercepts for each cluster.

In some cases, the timing and location of confounding factors are known in each cluster and their effects can be modeled. Often, investigators are unaware of all factors affecting the outcome. Throughout this paper we assume exposure to these factors are unknown. By incorporating only a fixed effect for time, the standard Hussey and Hughes model requires that the effects of time are common to all clusters, and that the correlation between any two observations in the same cluster are independent of time.

### 2.2. Random period models eq under confounding due to external factors

Since misspecification of the time effects can lead to biased estimates of the intervention effect and its standard error, it has been recommended to incorporate a random cluster-by-time period interaction effect.^13, 17, 20^ This formulation, referred to as the Hooper/Girling model,^21, 22^ allows the random intercept for each cluster to vary by time period. This model is discussed in Section 2.2.1 below.

#### 2.2.1. Random cluster-by-discrete time effect (uncorrelated, with single variance)

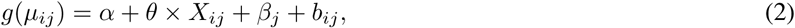

where 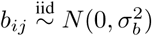 are the time-varying random intercepts for each cluster *i* at time period *j, i* = 1, …, *N* and *j* = 1, …, *n*. These cluster-specific random effects are intended to capture the effects of external factors on the outcome by allowing unique secular trends for each cluster.

There are limitations to the model above that arise from the distributional assumptions regarding the random effects *b*_*ij*_: Constant variance over time, independence of the random effects, and identical correlation between two observations in the same cluster between any two time periods. If exposure to confounding factors increases over time, the variance of the random effect may increase as well. The assumption of independence of the random effects may be violated in the scenario where confounding factors impact clusters at all time intervals after exposure. Such a scenario could lead to greater similarity of random effects during time intervals that are entirely before or entirely after exposure than for random effects for a mix of intervals that are both before and after exposure. Furthermore, while the Hooper/Girling model allows both within-cluster and within-period variation, it requires that the correlation between two observations in the same cluster be identical within and across two time intervals.

#### 2.2.2. Random cluster-by-discrete time effect (unstructured covariance)

To account for the limitations of model 2, we propose an unstructured covariance for the random cluster-by-discrete time interaction terms:

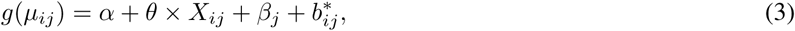

where 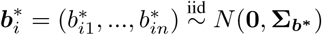 for *i* = 1, …, *N*. Similar to model 2, the cluster-specific random effects 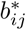 allow unique secular trends for each cluster. However, the unstructured covariance matrix 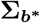 imposes no restrictions on the variance across time periods nor on the correlation between the latent time effects within each cluster, albeit with the assumption that the correlation structure is the same for all clusters. While model 3 imposes less restrictions than model 2, it requires the estimation of 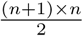 covariance parameters, which may greatly reduce the power to detect an intervention effect and may lead to issues of identifiability of regression parameter estimates.

#### 2.2.3 Random cluster by linear time effect

An alternative to models 2 and 3 is to include a random slope for time in each cluster:

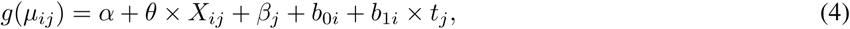

where 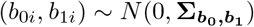and 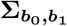 is the 2 *×* 2 covariance matrix for (*b*_0*i*_, *b*_1*i*_), *i* = 1, …, *N*, and *t*_*j*_ is the time between the study start date and the beginning of time period *j*. The random effects *b*_1*i*_ allow for unique linear time trends in each cluster. Model 4 assumes the variance in the outcome increases with time and imposes restrictions on the correlation between observations within a cluster, and is therefore more restrictive than model 3.

### 2.3. Random period models under confounding due to external factors and early adoption

We consider the scenario where control clusters, unbeknownst to the investigator, prematurely adopt components of the intervention before the scheduled roll-out time. We refer to this scenario as early adoption. In SWDs, there are a greater number of control clusters at risk for early adoption in the beginning of the study compared to later on. In addition, the probability that a control cluster prematurely adopts aspects of the intervention may change over time. Because exposure to early adoption is dependent on calendar time, this can be viewed as a special case of confounding by time in the control clusters only.

The models described in Section 2.2 are insufficient for this setting where confounding differentially impacts control and intervention clusters. To account for this scenario, such as early adoption by control communities, we consider group*×* time interaction models which incorporate distinct fixed and random time effects for control clusters. Unlike group*×*time interaction models for time-varying treatments, these models are intended to account for situations where secular trends systematically differ between intervention and control clusters.

In models 5 through 7, we incorporate a fixed discrete time effect, *β*_*j*_, that is common to all clusters. Because incorporating a group-by-discrete time interaction may lead to loss of power and issues of identifiability, we assume a fixed linear time effect in the control group by incorporating the term 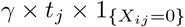 in models 5 through 7. In this setting, incorporation of a linear time effect in the control group assumes that early adoption of a beneficial intervention causes overdose death rates to monotonically decrease with time. We discuss these models in Sections 2.3.1 through 2.3.3.

#### 2.3.1. Random cluster-by-intervention-by-discrete time effect (uncorrelated, with single variance for each group)

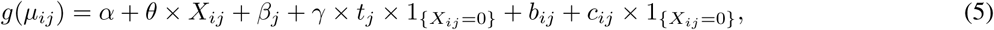

where *γ × t*_*j*_ is the difference in the time effect between control and intervention clusters at time period *j*, and 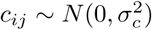 is a random time effect for control cluster *i* at time period *j* for *i* = 1, …, *N* and 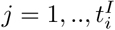, where 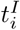 is the time period in which the intervention was scheduled to be rolled-out to cluster *i*. The (control) cluster-specific random effects, *c*_*ij*_, are intended to capture the effects of premature exposure to intervention components by control clusters. Control clusters that prematurely adopt intervention components are therefore expected to have larger *c*_*ij*_ (in magnitude). Model 5 is an extension to Model 2 (Hooper/Girling), in which the variance of the cluster-by-time period random effects systematically differ between intervention and control clusters. This model is subject similar limitations as those discussed in Section 2.2.1.

#### 2.3.2. Random cluster by-intervention-by-discrete time effect (unstructured covariance for each group)

To account for the limitations of model 5, we propose allowing an unstructured covariance for the random cluster-by-discrete time interaction terms in each group:

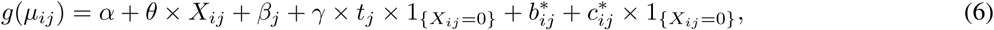

where 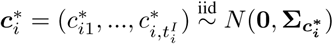 for *i* = 1, …, *N*. Here 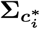 consists of the first 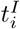 rows and columns of 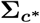, where 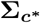 is the unstructured covariance matrix for a control cluster scheduled to receive the intervention during the final period of the roll-out phase. Similar to model (5), the cluster-specific random effects,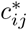, are intended to capture the effects of premature exposure to intervention components. This is an extension of model 3, which assumes the random cluster-by-discrete time effects shared a common (unstructured) covariance matrix. While the random effect covariance structure in model 6 has less restrictions than in model 5, it requires the estimation of 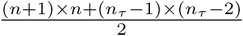 covariance parameters and is subject to similar limitations as discussed in Section 2.2.2. Here we define *n*_*τ*_ as the number of time periods prior to the follow-up phase of the study.

#### 2.3.3. Random cluster by-intervention-by-linear time effect

Similar to model 4, we can impose a random slope for time in each group.

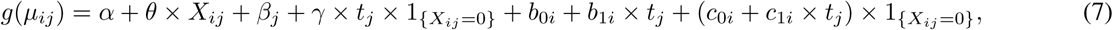

where 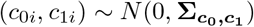 and 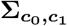 is the 2 *×* 2 covariance matrix for (*c*_0*i*_, *c*_1*i*_), *i* = 1, …, *N*. The cluster-specific random effects *c*_1*i*_ allow for unique linear time trends in each control cluster and are intended to capture the effects of premature adoption of intervention components.

#### 2.3.4. Fixed effects for linear time in all clusters

To further limit loss of power and potential issues of identifiability due to estimation of too many parameters, we replace the fixed discrete time effect in models 5 through 7, *β*_*j*_, with a fixed linear time effect, *β × t*_*j*_, in models 8 through 10. This strategy may be useful in situations when the combined effects of confounding factors result in monotonic changes in the outcome over time. For example, the implementation of a new policy intended to reduce opioid prescriptions in conjunction with media awareness campaigns may decrease the level of opioid overdose deaths with time.

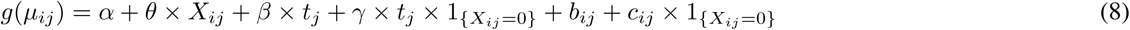

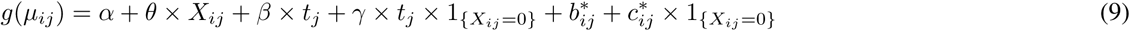

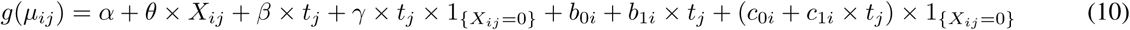

### 2.4. Estimation

These models can be fit by specifying the Poisson family in the *glmer* function (package: *lme4*) in ***R***.^25^ Of note, when the outcome is a summary measure for each cluster (as is the case in our motivating example), we ran into estimation difficulties when specifying random cluster-by-discrete time effects when the outcome distribution was assumed to be Gaussian. This is due to incorporation of a residual term for each cluster for each time period, which yields unidentifiable random effects. This issue does not arise under Poisson distributional assumptions, since the variance is directly proportional to the mean and thus residual terms are not estimated. To accommodate the Gaussian distribution assumption in this scenario, multiple communities per cluster^13, 17, 20^ or multiple intervals per time period would be needed.

It is important to distinguish between the impact of calendar time on the intervention assignment effect and on the intervention effect itself. Because of randomization, the intervention assignment effect is not confounded by calendar time; a randomized comparison will yield an unbiased estimate of the effect of assignment to the intervention initiation time. This effect includes all consequences of starting at that time, including the initiation of treatment. Nonetheless, our target of estimation, the effect of intervention alone, is confounded by calendar time.

In the models above, the fixed time effects *β*_*j*_ and *β* and the random time effects *b*_*ij*_,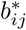, and *b*_1*i*_ are modeled to capture confounding factors that influence all clusters. The fixed time effects *γ* and random time effects *c*_*ij*_,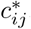, and *c*_1*i*_ are modeled to capture confounding factors that influence control communities only. These effects are all identifiable because the design matrices are full rank. Without additional assumptions, however, the above models cannot distinguish among the effects of external factors, early intervention adoption, or of other time-varying factors. Rather, they model temporal trends that may be a consequence of these factors; such models allow for assessment of the impact of such trends on bias, power, and Type 1 error. When used in analysis, our simulation studies demonstrate that these models can reduce bias in estimation of intervention effects, in the settings appropriate for their use.

## 3. Simulation study

We conduct a simulation study to investigate the impact of confounding factors on the bias, coverage probability of 95% confidence intervals, power, and Type 1 error of the intervention effect estimate. The data are simulated based on the motivating example described in Section 1.1. In this setting, we have *N* = 18 clusters and *n* = 13 time periods. The intervention is rolled out to each cluster according to the time line provided in Figure 1. In all data generating models, the outcomes *Y*_*ij*_ are simulated from a Poisson distribution, and represent the number of opioid overdose deaths in cluster *i* during time period *j*. Here *g* is the log link, and the population size of cluster *i, O*_*i*_, is included as an offset in all models. That is, *g*(*µ*_*ij*_) = *log*(*E*[*Y*_*ij*_]*/O*_*i*_). The intercept *α* is set to -10 and the standard deviation of the random intercept,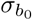, is set to 0.30. These numbers were determined using the opioid death counts in the 18 South Carolina clusters between 2016 and 2018. The data are provided in Supplementary Table 1. The intervention effect *θ* is set to log(0.6), which represents the target 40% reduction in opioid overdose deaths due to intervention (Section 1.1).

### 3.1. Simulation scenarios

We apply models 1 through 10 under four general scenarios: (1) standard, (2) presence of external factors, (3) early intervention adoption, and (4) presence of external factors and early adoption. The data generation process for each scenario is summarized in Table 1, and is described in more detail below.

**Table 1.**
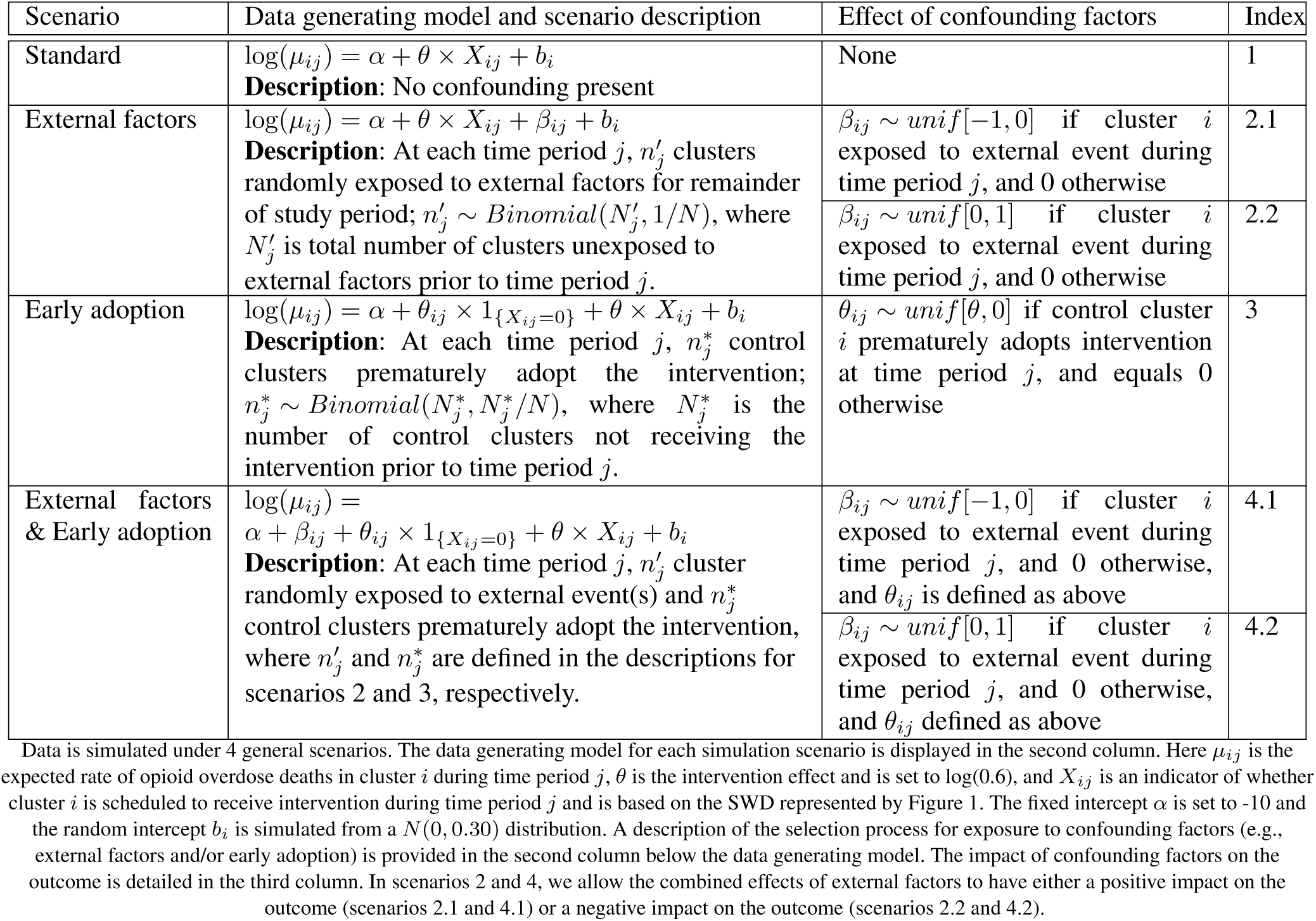
Simulation scenarios.

| Scenario | Data generating model and scenario description | Effect of confounding factors | Index |
| --- | --- | --- | --- |
| Standard | $\log(\mu_{ij}) = \alpha + \theta \times X_{ij} + b_i$<br>**Description:** No confounding present | None | 1 |
| External factors | $\log(\mu_{ij}) = \alpha + \theta \times X_{ij} + \beta_{ij} + b_i$<br>**Description:** At each time period $j$, $n'_j$ clusters randomly exposed to external factors for remainder of study period; $n'_j \sim \text{Binomial}(N'_j, 1/N)$, where $N'_j$ is total number of clusters unexposed to external factors prior to time period $j$. | $\beta_{ij} \sim \text{unif}[-1, 0]$ if cluster $i$ exposed to external event during time period $j$, and 0 otherwise | 2.1 |
| | | $\beta_{ij} \sim \text{unif}[0, 1]$ if cluster $i$ exposed to external event during time period $j$, and 0 otherwise | 2.2 |
| Early adoption | $\log(\mu_{ij}) = \alpha + \theta_{ij} \times 1_{\{X_{ij}=0\}} + \theta \times X_{ij} + b_i$<br>**Description:** At each time period $j$, $n_j^*$ control clusters prematurely adopt the intervention; $n_j^* \sim \text{Binomial}(N_j^*, N_j^*/N)$, where $N_j^*$ is the number of control clusters not receiving the intervention prior to time period $j$. | $\theta_{ij} \sim \text{unif}[\theta, 0]$ if control cluster $i$ prematurely adopts intervention at time period $j$, and equals 0 otherwise | 3 |
| External factors & Early adoption | $\log(\mu_{ij}) = \alpha + \beta_{ij} + \theta_{ij} \times 1_{\{X_{ij}=0\}} + \theta \times X_{ij} + b_i$<br>**Description:** At each time period $j$, $n'_j$ cluster randomly exposed to external event(s) and $n_j^*$ control clusters prematurely adopt the intervention, where $n'_j$ and $n_j^*$ are defined in the descriptions for scenarios 2 and 3, respectively. | $\beta_{ij} \sim \text{unif}[-1, 0]$ if cluster $i$ exposed to external event during time period $j$, and 0 otherwise, and $\theta_{ij}$ is defined as above | 4.1 |
| | | $\beta_{ij} \sim \text{unif}[0, 1]$ if cluster $i$ exposed to external event during time period $j$, and 0 otherwise, and $\theta_{ij}$ defined as above | 4.2 |

In the first scenario (standard) we assume no confounding is present; all clusters receive the intervention during the scheduled time period and no external factors influence the outcome. The data are generated according to scenario 1 in Table 1. In the second scenario, all clusters not currently exposed to external factors by time period *j* have a 1 in N chance of exposure. Once exposed, each cluster continues to be exposed through the remainder of the study period. This is intended to reflect the situation where external factors have long-lasting effects, such as new public policies limiting opioid prescriptions and media awareness campaigns. The number of clusters exposed to external factors at each time period *j*, 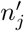, is simulated from a *Binomial*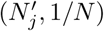 distribution, where 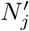 is the total number of clusters unexposed to the external factors prior to time period *j*.

Once exposed, the effect of external factors on the outcome in cluster *i* is set to vary uniformly at each time period *j*. Specifically, we let 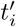 denote the time period in which cluster *i* is exposed to external factors. We simulate *β*_*ij*_ under 2 settings. For 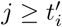, we simulate *β*_*ij*_ *∼ unif* [*−*1, 0], corresponding to a positive effect of external factors on the outcome opioid overdose deaths (scenario 2.1). This is intended to represent a rising tide of events aimed at improving outcomes. We set *β*_*ij*_ = 0 for 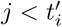, indicating that cluster *i* has yet to be exposed to external factors. To represent a negative impact of external factors on the outcome, we simulate *β*_*ij*_ *∼ unif* [0, 1] for 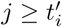 and *β*_*ij*_ = 0 for 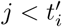 (scenario 2.2).

In the third scenario (early adoption), the number of clusters prematurely adopting the intervention during each time period *j*, 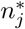, is simulated from a *Binomial* 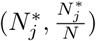distribution. Here 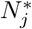 is the number of control clusters which have not adopted any intervention components by time period *j*. This set up reflects the nature of SWD, where the number of control clusters at risk for early adoption decreases with time as more clusters crossover to the intervention group by design. However, the probably of exposure to early adoption for a control cluster likely increases with time as interventions become more widely available. Once exposed to early adoption, we assume that exposure continues until the scheduled roll-out of the intervention, at which point the cluster receives the full intervention effect.

For all time periods *j* in which a cluster is subjected to early adoption, the magnitude of the early intervention adoption effect is set to vary uniformly. Denote 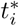as the time period of early adoption for cluster *i* and denote 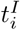 as the time period in which the intervention was scheduled to be rolled-out to cluster *i*. We simulate *θ*_*ij*_ *∼ unif* [*θ*, 0] for 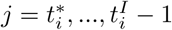, and *θ*_*ij*_ = 0 otherwise. Thus the maximum effect of early intervention adoption on the outcome for exposed control clusters is set to *θ*. In this scenario, the confounding factor (early adoption) has a positive effect on the outcome in the control group only.

In the fourth scenario, the effects of external factors and early adoption are simulated in the same manner as in scenario 2 and scenario 3, respectively. In this scenario, the data are generated according to scenarios 4.1, where external factors have a positive effect on the outcome, and scenario 4.2, where external factors have a negative effect on the outcome. Additional detail is provided in Table 1. We note that in the latter scenario (4.2), the negative impact of external factors on the outcome is partially offset by the positive impact of early adoption on the outcome.

### 3.2. Results

Simulation results in are presented in Table 2. Under scenario 1 (standard), all models yield unbiased estimates of the intervention effect, reach nominal confidence interval coverage rates, and preserve Type 1 error. Model 1, which does not include any random effects for time, yields unbiased estimates of the intervention effect in scenario 2 (external factors), where early adoption is not present and therefore confounding does not differentially impact control and intervention clusters. However, coverage probabilities are below the nominal level of 0.95 and Type 1 error is inflated. Model 1 is heavily biased in scenarios 3 (early adoption) and 4 (external factors & early adoption), where confounding differentially impacts intervention and control clusters due to the early adoption.

**Table 2.**
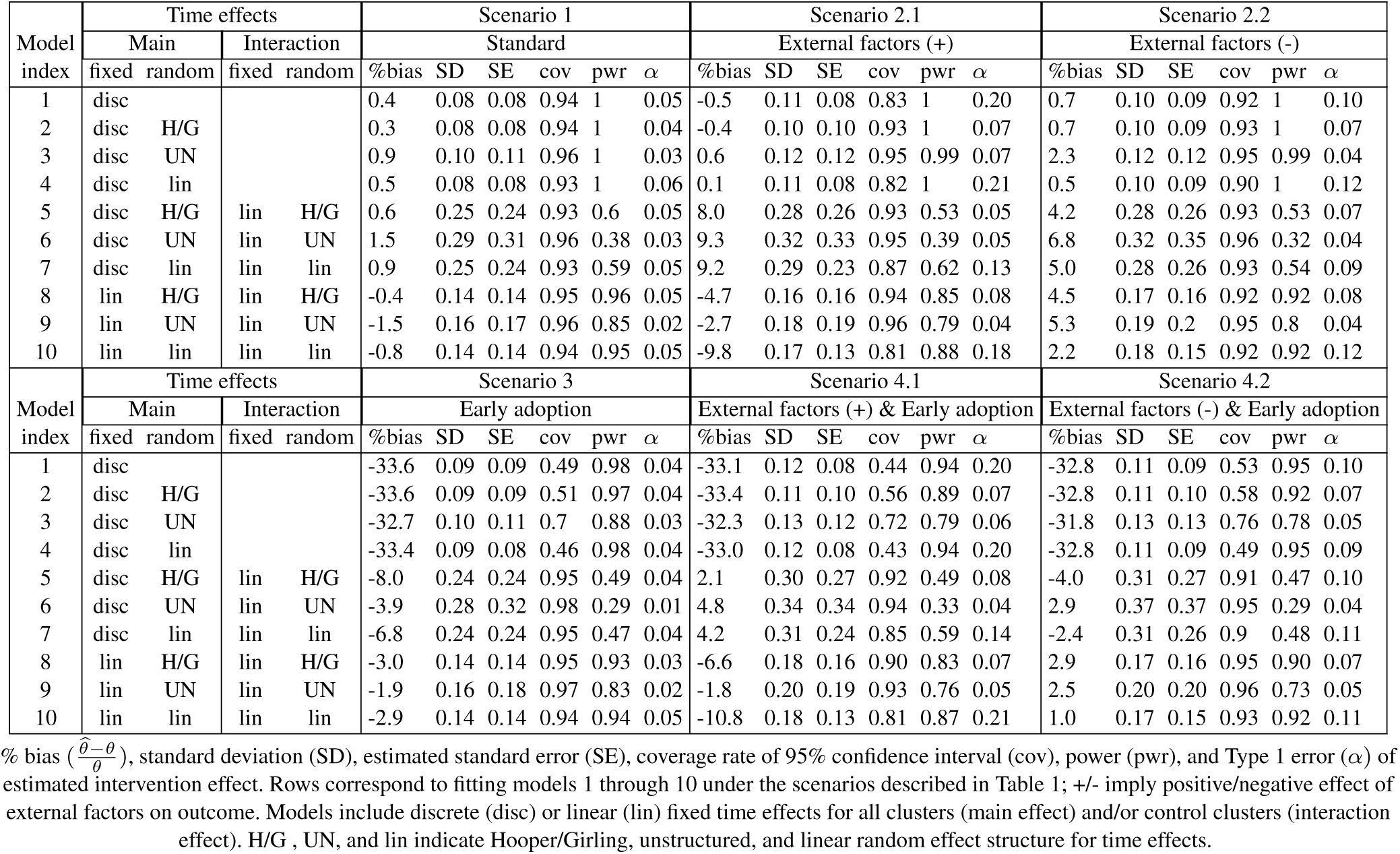
Simulation Results.

Models 2 through 4, which include random cluster-by-time effects, yield unbiased estimates of the intervention effect when confounding does not differentially impact control and intervention clusters (i.e., scenarios 1 and 2). Model 3, which assumes an unstructured covariance for the random cluster-by-time interactions, performs the best with regard to coverage probabilities of the 95% confidence intervals and Type 1 error preservation. Model 2, which assume a single variance for the random cluster-by-time interactions (Hooper/Girling model), perform slightly worse on these metrics. Model 4, which assume a random slope for time for each cluster, has the highest inflation in Type 1 error with coverage probabilities well below the nominal level of 0.95. Models 2 through 4 are overpowered for scenarios 1 and 2. These models perform poorly when early adoption is present in scenarios 3 and 4. The estimated intervention effect is reduced by 31.8% to 33.6% in these scenarios.

The performance of models with and without intervention-by-time interactions is compared in Figure 2. Models without intervention-by-time interactions are displayed in the left column, where the blue shapes correspond to models 2 through For comparison, we include models which replace the discrete main effect for (fixed) time in models 2 through 4, *β*_*j*_, with the linear fixed effect *β × t*_*j*_. These models are labeled by the red shapes in the left column of Figure 2. The models in the right column of Figure 2 include fixed and random intervention-by-time interactions. These models correspond to models 5 through 10.

**Figure 2.**
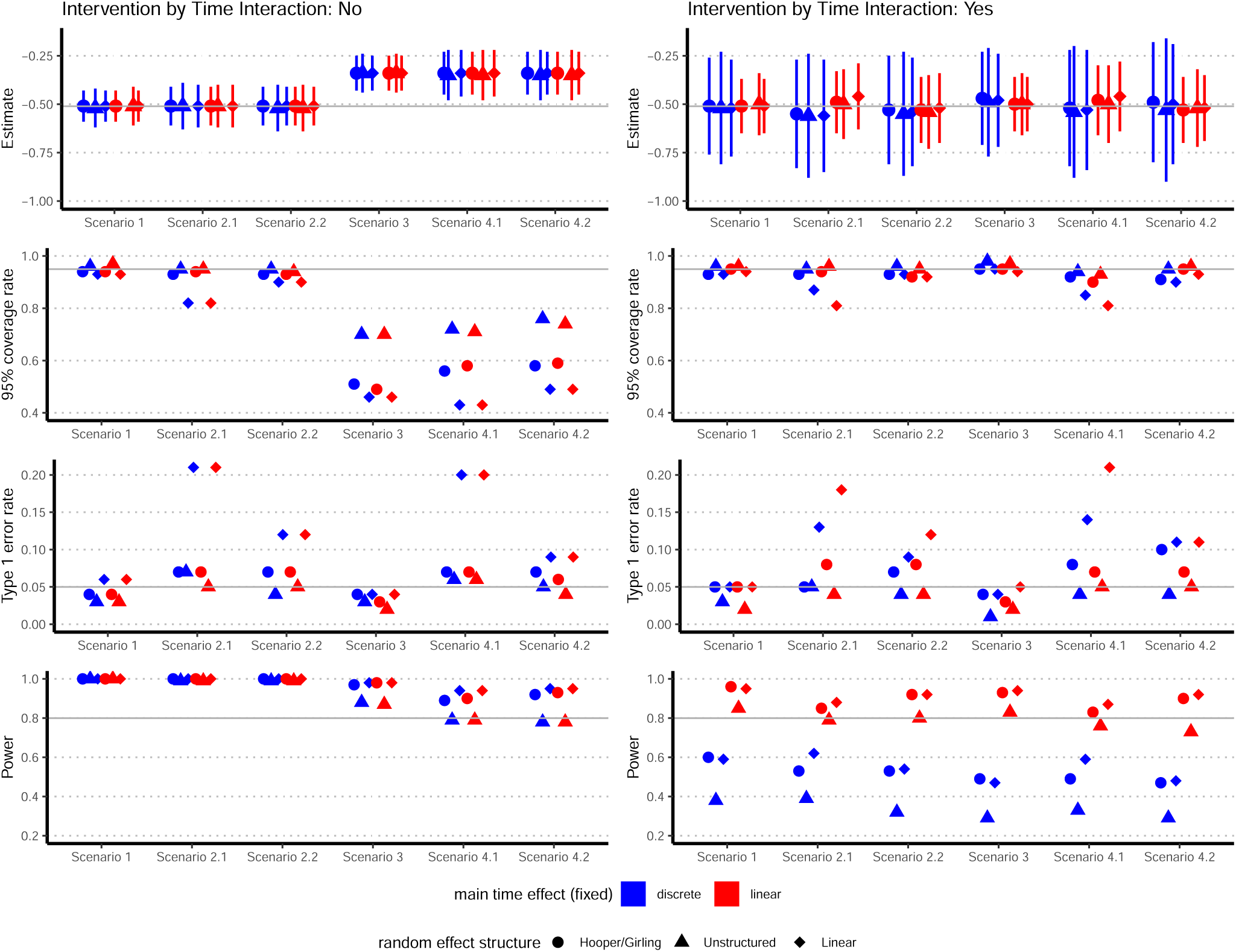
Performance of models with and without fixed and random intervention-by-time interactions. Models are compared across scenarios listed in Table 1. First row compares intervention effect estimates *±* empirical standard error. Horizontal gray line: true intervention effect *θ* = log(0.6). Second, third, and fourth rows compare models on coverage rate of 95% confidence intervals, Type 1 error rate, and power, respectively. Horizontal gray lines indicate 95% coverage rate, 0.05 Type 1 error rate, and a power of 0.80 in the second, third, and fourth rows, respectively. Covariance structure for cluster-by-time random effects: Hooper/Girling labeled by circles, unstructured labeled by triangles, and linear labeled by diamonds. Models which incorporate a discrete term for the main effect for time (fixed) are labeled by blue shapes; models which incorporate a linear term are labeled by red shapes. Models without intervention-by-time interactions displayed in left column, where blue shapes correspond to models 2 through 4 in Section 2. Models with intervention-by-time interactions displayed in right column, and correspond to models 5 through 10 in Section 2. Models 5 through 10 include a linear time effect for the fixed intervention-by-time interaction term.

Models 5 through 10 perform well in all simulation scenarios. When no confounding or early adoption is present (scenario 1), these models yield little bias, reach nominal coverage probabilities for 95% confidence intervals, and preserve Type 1 error. When only confounding due to external factors is present (scenario 2), all models have less than 10% bias. These models generally reach nominal confidence interval coverage probabilities and preserve Type 1 error. The exceptions are models 7 and 10, which include random cluster-by-group-by-linear time interaction effects.

When confounding differentially impacts intervention and control clusters due to early adoption (scenarios 3 and 4), models 5 through 10 greatly reduce the bias in the intervention effect estimate compared to models which do not include intervention-by-time interactions. Models 6 and 9, which assume an unstructured covariance for the random cluster-by-group-by-discrete time interactions, generally yield the lowest bias, while reaching nominal coverage probabilities of the 95% confidence intervals and preserving Type 1 error. Models 5 and 8 (Hooper/Girling models) perform slightly worse on these metrics. Models 7 and 10 have the highest inflation in Type 1 error and coverage probabilities well below the nominal level.

The choice of a discrete or linear term for the fixed time effect does not impact power in models without intervention-by-time interactions. This result is expected given the findings of Grantham et al.^26^ For models which include intervention-by-time interaction terms (i.e., models 5 through 10), the strongest correlate of power is the fixed effect for time. Models with discrete time effects, which incorporate a parameter for each time period, have much lower power compared to models which incorporate a single parameter corresponding to a linear time effect. Models with a random slope for time achieve the highest power (models 7 and 10), while models which assume an unstructured covariance for random cluster-by-time effects achieve the lowest power (models 6 and 9).

## 4. Discussion

SWDs and alternative cluster randomized trials are currently being used to implement interventions to reduce opioid overdose deaths in communities across several states. However, these interventions are competing with newly initiated public policies and media awareness campaigns. Furthermore, control communities may adopt components of proposed interventions as they become readily available. These scenarios induce confounding by time. In this paper, we have demonstrated the consequences of inadequately accounting for time effects in these scenarios. Given the difficulties in capturing the timing and exposure levels of such confounding factors, we considered mixed-effects models which incorporate random effects to capture these latent confounding factors. We discussed the limitations of commonly used models in the context of proposed SWDs to combat the opioid epidemic, and proposed solutions to accommodate deviations from these assumptions.

Through our simulation study, we showed that recently proposed mixed effects models are sensitive to the type of factors which induce confounding by time. The Hooper/Girling model^17, 20^ offers an improvement over the standard Hussey and Hughes model,^16^ but may result in severely biased estimates of the intervention effect when secular trends systematically differ between intervention and control clusters. Even in scenarios where these models do capture the secular trend, we demonstrated how incorrect specification of the cluster-level covariance over time can yield under coverage of confidence intervals and inflation of Type 1 error. Similar conclusions have been reached in other studies.^3, 22, 22, 27, 28^

Alternatively, models which allow secular trends to systematically differ between the intervention and control clusters through incorporation of fixed and random group-by-time effects offer a major improvement in terms of bias reduction. Our simulation studies confirmed that models incorporating unstructured cluster-level covariances for the random intervention-by-cluster-by-time interaction terms yielded nominal confidence interval coverage rates and preserved Type 1 error (i.e., models 6 and 9). However, these models may be underpowered for certain parameterizations of the fixed time effects.

The proposed mixed-effects modeling framework discussed in this paper treats external factors and early intervention adoption as latent processes; these confounding factors are accounted for through the incorporation of random effects. This modeling strategy is useful in practice because investigators are often unaware of all confounding factors affecting the outcome, let alone each cluster’s level of exposure to these factors. Furthermore, as evidenced by the results of our simulation study, these models have a certain level of robustness to the assumed distribution of the random effects.

### 4.1. Limitations and Future Research

An important limitation of the models we discussed in this paper is that they assume the intervention effect does not vary with time. Models attempting to account for both secular trends which differ by intervention group, and time-varying treatment effects, may lead to unidentifiable intervention effect estimates. Furthermore, the approach discussed here is limited to GLMM. Generalized estimating equations (GEE) also allow for correlation in the outcomes, and are robust to misspecification of the covariance structure.^29^ Ren et al. show that GEE is more robust to model misspecification than linear mixed models when the random intercepts differ by intervention group. Future work is needed to explore the performance of GEE models in the context considered here, where secular trends in the control and intervention clusters arise from different mechanisms, and differentially impact clusters within each group. Several nonparametric methods have also been proposed that use within-period^3, 27, 30^ or between period^3^ comparisons to account for confounding by time. While these models are robust to misspecification of the random effects, the performance of these models when confounding factors differentially impact intervention and control clusters, such as in the case of early intervention adoption, has not been explored.

Although our paper focused on stepped-wedge designs, the secular trends in outcomes induced by rising tides and early intervention adoption may also be present in other types of cluster randomized trials. Simulation studies are needed to determine the impact of external factors and early adoption on the bias of the intervention effect estimate, Type 1 error, and power in these settings. Grantham et al. establish a sufficient condition for when the choice of time parameterization does not impact the variance of the estimated intervention effect in cluster randomized trials.^26^ This information can be useful in the planning of such trials. While Grantham et al establish that categorical or linear fixed time effects do not impact the variance estimator of the intervention effect in SWD, this was not the case when group-by-time interaction terms were modeled (as demonstrated by our simulation study). Future investigation is needed to establish sufficient conditions in the presence of interactions.

### 4.2. Conclusion and Recommendations

Stepped-wedge designs are particularly suitable for epidemics and pandemics, where complex health interventions are needed as soon as possible. Given the urgency of such situations, other policies and interventions aimed at improving outcomes will likely be implemented in concurrence with any proposed interventions. This induces confounding by time, which we have shown can severely bias the intervention effect estimate in SWDs when not properly accounted for. Studies must therefore consider these scenarios during the planning stages. We have shown that models which incorporate fixed and random group-by-time effects are effective in reducing bias when confounding factors differentially impact intervention and control clusters. However, incorporation of such time effects may impact power and must be accounted for in sample size calculations. Models incorporating an unstructured covariance for the random intervention-by-cluster-by-time interaction effects are most effective in reducing bias, reaching nominal confidence interval coverage, and preserving Type 1 error; but they can lead to severely under powered studies when used in conjunction with discrete fixed time effects. One strategy is to use fixed parametric time effects and an unstructured covariance; these models perform reasonably well in the scenarios considered here. Alternatively, one can impose a more restrictive covariance structure such as exponential decay over time, Hooper/Girling covariance, or random slopes for time. To account for potential under coverage of confidence intervals and Type 1 error inflation, randomization based inference should be used.^31, 32^ Simulation studies may be needed to estimate sample size and power for certain models, as formulas to calculate them are not currently available for many random effect covariance structures.

## Data Availability

The dataset supporting the conclusions of this article is included within the web appendix of this article. We are making the computer code available. Once accepted, our code will be deposited to github, and will be linked to the final published article.

## 5. Declarations

## Acknowledgments

Not applicable.

## Authors’ contributions

LR, MH, and VG conceived the methods and LR drafted the manuscript. VDG and MH helped write the paper. VDG, MH, and AHL critically reviewed the contents of the paper. AHL designed the study used as a motivating example in this paper. LR carried out the simulation study.

## Consent for publication

Not applicable.

## Ethics approval and consent to participate

No ethical approval or consent was required for this simulation study based on a proposed trial design.

